# Deep kinetoplast genome analyses result in a novel molecular assay for detecting *Trypanosoma brucei gambiense*-specific minicircles

**DOI:** 10.1101/2022.03.15.22272384

**Authors:** Manon Geerts, Zihao Chen, Nicolas Bebronne, Nicholas J. Savill, Achim Schnaufer, Philippe Büscher, Nick Van Reet, Frederik Van den Broeck

**Author notes:** Corresponding author: Frederik Van den Broeck. Co-senior authors: Frederik Van den Broeck and Nick Van Reet.

## Abstract

The World Health Organization targeted *Trypanosoma brucei gambiense* (*Tbg*) human African trypanosomiasis for elimination of transmission by 2030. Sensitive molecular markers that specifically detect *Tbg* type 1 (*Tbg1)* parasites will be important tools to assist in reaching this goal. Here, we aim at improving molecular diagnosis of *Tbg*1 infections by targeting the abundant mitochondrial minicircles within the kinetoplast of *Trypanosoma brucei* parasites. Using Next-Generation Sequencing of total cellular DNA extracts, we assembled and annotated the kinetoplast genome and investigated minicircle sequence diversity in 38 animal- and human-infective trypanosome strains. Computational analyses recognized a total of 241 Minicircle Sequence Classes as *Tbg*1-specific, of which three were shared by the 18 studied *Tbg*1 strains. We then developed a novel multiplex quantitative PCR assay (*g*-qPCR3) targeting one *Tbg*1-specific minicircle and three *Tbg*1-specific or Trypanozoon-specific markers. Molecular analyses revealed that the minicircle-based assay is applicable on animals and is as specific as the *TgsGP*-based assay, the current golden standard for molecular detection of *Tbg*1. The median copy number of the targeted minicircle was equal to eight, suggesting that our minicircle-based assay may be used for the sensitive detection of *Tbg*1 parasites. Finally, annotation of the targeted minicircle sequence indicated that it encodes genes essential for the survival of the parasite, and will thus likely be preserved in natural *Tbg*1 populations. These results demonstrated that our minicircle-based assay is a promising new tool for reliable and sensitive detection of *Tbg*1 infections in humans and animals.

## INTRODUCTION

Human African trypanosomiasis (HAT), also known as sleeping sickness, is a vector-borne disease caused by two *Trypanosoma brucei* (*Tb*) subspecies and transmitted by tsetse flies. *Trypanosoma brucei rhodesiense* (*Tbr*) causes acute infections in East Africa, whereas *Trypanosoma brucei gambiense* (*Tbg*) causes chronic infections in West and Central Africa (1). *Trypanosoma brucei rhodesiense* and *Tbg* type I (*Tbg*1) are defined by the presence of truncated variant surface glycoprotein (VSG) genes, respectively the serum-resistance-associated (SRA) gene (2) and the *Tbg*-specific glycoprotein (TgsGP) gene (3) that play a role in infectivity to humans (4). Unlike *Tbr, Tbg*1 parasites are genetically homogeneous and form a monophyletic group (5, 6). They are responsible for the vast majority of the HAT-cases (85% of the 663 newly reported HAT-cases in 2020) (7). Infections by a third group of human-infective trypanosomes - *Tbg* type II (*Tbg*2) that lack the SRA and TgsGP genes - are extremely rare (8). Like the non-human-infective *T. b. brucei (Tbb), Tbg*2 is genetically and phenotypically highly diverse (8).

The WHO targeted *Tbg*-HAT (gHAT) for elimination as a public health problem by 2020 and for global elimination of transmission (EOT) to humans (i.e. zero reported cases) by 2030 (9). Elimination of gHAT as a public health problem has been reached in several countries and HAT foci, and recently Togo and Côte d’Ivoire have been validated as such by WHO (10, 11). However, EOT remains challenging due to imperfect diagnostics and the risk of re-emergence from asymptomatic human infections and/or a possible animal reservoir (12– 14). Current serological tests based on the *Tbg*-specific LiTat 1.3 and LiTat 1.5 VSG antigens (15–20) still require validation in different animal species. The golden standard for molecular detection of *Tbg*1 involves the single-copy *TgsGP* gene (21–23), but the analytical sensitivity of these tests is limited because they target a hemizygous single-copy gene (24). Among the molecular tests, several studies have proposed various genotyping techniques such as isoenzymes, ribosomal genes, VSGs, SNPs and microsatellites (25–32), but these have some disadvantages such as the requirement of multiple PCR reactions or high amounts of input material, without necessarily increasing the sensitivity of *Tbg1* detection. To this end, alternative genetic markers are much needed to reliably and with improved sensitivity demonstrate *Tbg*1 infection in humans and in animals, including the tsetse fly vector.

Previous reports indicated that *Tbg1*-specific minicircle sequences exist in the kinetoplast DNA (33, 34). The kinetoplast DNA (kDNA), unique to the single mitochondrion of unicellular flagellates of the order Kinetoplastida, is a giant network of dozens of homogenous maxicircles (20-30 kb) interlaced with hundreds to thousands of heterogeneous minicircles (0.5-2.5 kb) (35). Maxicircles are homologous to the mitochondrial genome of other eukaryotes and encode components of the respiratory chain complexes and the mitoribosome. Minicircles generally consist of a ∼100 bp conserved sequence region that contains hyper conserved sequences named Conserved Sequence Blocks (CSBs), and a variable region including genes encoding guide RNAs (gRNAs) that are responsible for directing post-transcriptional modification of the maxicircle-encoded messenger RNAs (35). A complete assembly and annotation of the kinetoplast genome of a lab-adapted *Tbb* strain identified 391 minicircle classes, encoding ∼1,000 gRNA genes (36). Analysis of kDNA minicircles is already used for *Leishmania* detection and differentiation (37, 38) and allows subtypes of *Trypanosoma evansi* to be distinguished (39). It has also been proposed for sensitive and specific detection of *Tbg*1 infection in humans (34) and animals (33). However, the exact nature of these sequences is unknown and their use for *Tbg1* diagnosis is cumbersome, requiring both nested PCR and DNA hybridization.

Recent advances in the development of bioinformatic tools now facilitate the assembly and annotation of the structurally complex kinetoplast genome (36, 40), allowing us to investigate minicircle sequence diversity in tens to hundreds of samples simultaneously (41). Therefore, we used next-generation sequencing of total cellular DNA extracts as a strategy to investigate minicircle sequence diversity in 38 animal- and human-infective trypanosome strains from diverse geographical origins. Following a series of computational analyses, we identify minicircles that were present in all *Tbg*1 strains and absent in all *Tbb, Tbg2* and *Tbr* strains. Using a newly developed quantitative PCR (qPCR), we demonstrate that our minicircle-based assay reliably identifies *Tbg1* within the Trypanozoon subgenus. Furthermore, we show that the copy number of the targeted minicircle was on average 4-fold higher compared to the hemizygous single-copy *TgsGP* gene, suggesting that our minicircle-based assay may be used for sensitive detection of *Tbg*1 infections in humans and animals.

## METHODS

### Ethics statement

Expansion of bloodstream form trypanosome populations in mice received approval from the Animal Ethics Committee of the Institute of Tropical Medicine (DPU2017-1).

### DNA extraction and sequencing

To cover a wide geographical range, we included a total of 18 *Tbg*1 strains isolated from humans between 1952 and 2008 from Cameroon (n = 2), the Democratic Republic of the Congo (n = 10), the Republic of Congo (n = 2), Côte d’Ivoire (n = 3) and South Sudan (n = 1) (Supplementary Table 1). For comparative purposes, we also included nine *Tbb*, five *Tbg*2 and six *Tbr* strains (Supplementary Table 1).

All 38 strains were propagated as bloodstream form populations in OF1 mice (Charles River, Belgium) and purified from the infected mouse blood via DEAE ion exchange chromatography (42). Purified trypanosomes were sedimented by centrifugation (17,000 g, 10 min at 4°C). DNA of 50 µL pure trypanosome sediment was extracted using the standard phenol:chloroform method (43), aliquoted at 1 ng/µl and stored at -20°C. The concentration of extracted DNA was determined using a Qubit 4 Fluorometer (Invitrogen by Thermo Fisher Scientific). Paired-end 150 bp sequences were generated using the DNA nanoball sequencing technology (DNBSEQ™) at the Beijing Genomics Institute (BGI) in Hongkong, China.

### Genomic analyses

Paired-end reads were aligned against the *Tbb* TREU927 v4.6 reference genome (available on https://tritrypdb.org) using SMALT v0.7.6 (https://www.sanger.ac.uk/tool/smalt-0/). A hash index of the reference genome was built using *k*-mer words of length 13 that were sampled every other position in the genome. Mapping was done using an exhaustive search for alignments with a minimum identity threshold (-y) of 80% and a maximum insert size for paired reads of 1,500 bp.

Variant calling and filtering was performed using the Genome Analysis Toolkit (GATK) v4.1.4.1 (44). First, reads were assigned to a single read-group with *AddOrReplaceReadGroups* and duplicated reads were marked with *MarkDuplicates*. Variants were then called for each strain separately with *HaplotypeCaller* using default parameters. The resulting gVCF files for all strains were combined with *CombineGVCFs* to allow joint genotyping with *GenotypeGVCFs*. Single Nucleotide Polymorphisms (SNPs) were extracted from the resulting VCF file with *SelectVariants* and filtered with *VariantFiltration* using the following parameters: QUAL < 500, DP < 5, QD < 2.0, FS > 60.0, MQ < 40.0, MQRankSum < -12.5, ReadPosRankSum < -8.0, --cluster_window_size 10 and -- cluster_size 3. Finally, we used BCFtools v1.10.2 (45) to extract bi-allelic SNP sites that were called in all *Tb* strains. Using the resulting set of genome-wide SNPs, we reconstructed a phylogenetic network with SplitsTree v4 (46) to infer the ancestral relationship among the 38 *Tb* strains.

The species identity of the *Tbg*1 and *Tbr* strains was confirmed *in silico* by investigating the presence of the TgsGP and SRA genes, respectively. To this end, MEGAHIT v1.2.9 (47) was used for *de novo* assembly of genomes of all 38 *Tb* strains using default parameters. The presence of TgsGP and SRA genes in the assembled contigs was then confirmed through a local BLAST search (48) using publicly available nucleotide sequences of TgsGP (21) (NCBI accession number: FN555993) and SRA (2) (NCBI accession number: Z37159) with the following parameters: minimum 90% identity, minimum e-value of 0.0001 and minimum alignment length of 500bp.

### Assembly of the kinetoplast genome

Reads that did not align to the *Tbb* TREU927 nuclear reference genome were extracted using SAMtools v1.9 (45) and converted to FASTQ format using GATK *SamToFastq*. These unmapped reads were aligned against the 23 kb maxicircle sequence of *Tbb* Lister 427 (GenBank accession id M94286) using SMALT and the same parameters as described above except that the hash index was built with *k*-mer words of length six and the reads were mapped with a minimum identity threshold (-y) of 90% and a maximum insert size (-i) of 500. SNP calling was done using GATK as described above, and we extracted only those SNPs that passed the quality criteria (see above) and that were present within the maxicircle coding region (1.3-16.3 kb). Similar to the analyses above for the nuclear genome, a phylogenetic network analysis was done using maxicircle coding SNPs with SplitsTree.

Reads that did not align to the maxicircle sequence of *Tbb* Lister 427 were extracted from the alignment file as described above, and used for the assembly of minicircle contigs. Before assembly, sequence reads were trimmed for high quality with fastp v0.20.0 (49) using the following parameters: allow for a maximum of 10% of bases per read that have a phred-scaled base quality below 30, trim bases at either end of the read when their phred-scaled quality is below 30, move a sliding window of 10 bp from front to tail and cut the read once the average phred-scaled base quality drops below 30, and only retain reads with a minimum and maximum length of 100 bp and 155 bp after trimming, respectively. Using KOMICS v1.8 *assemble (40)*, trimmed reads were used for *de novo* assembly of contigs using a *k*-mer list of 99, 109 and 119, and putative minicircle contigs were extracted based on the highly conserved sequence block 3 (CSB3) dodecamer (GGGGTTG[G/A]TGTA) (36, 50) Five minicircles had a slight variation of CSB3 (one with GGGG<u>G</u>TGGTGTA found in *Tbg*2 strain FEO and four with GGGGTT<u>A</u>GTGTA found in *Tbg*1 strains 15BT-relapse, OUSOU, NDIMI and ROUPO-VAVOUA--80-MURAZ-14). These minicircles were also retained. KOMICS *circularize* was then used to identify circular minicircle contigs by searching for overlapping fragments at either end of each contig; when a minicircle contig was classified as circular, the overlapping fragment at the start of the contig was removed. Using KOMICS *polish*, all circular minicircle contigs were oriented by putting the conserved sequence block 1 (CSB1) (GGGCGT[T/G]C) (50) at the start of each contig. One minicircle had a slight variation of CSB1 (GGGCGTG<u>T</u> found in *Tbg*2 strain MSUS-CI-78-TSW-157), which was specified to KOMICS to allow proper reorientation for this minicircle. Finally, KOMICS *polish* was also used to remove duplicate sequences, which was achieved by extracting the representative sequences (cluster centroids) of all clusters identified at 97% identity with VSEARCH v2.14.2 (51).

The quality of the minicircle assembly was assessed by re-aligning the unmapped reads to the assembled minicircles using SMALT. Before mapping, we first extended the circularized minicircle sequences by copying the last 150 bp at the start of each sequence to minimize the number of clipped reads at either end of the assembled minicircles, using a custom <u>python script</u> implemented in KOMICS. Following mapping with SMALT with exhaustive search (-x) and a percent identity of 97% (-y), we have calculated the following metrics using a <u>bash script</u> implemented in KOMICS: number of reads, number of mapped reads, number of properly paired reads, number of reads with mapping quality >=20, number of CSB3-containing reads, number of mapped CSB3-containing reads and number of perfectly aligned CSB3-containing reads (i.e. alignments without any insertions or deletions). The proportion of perfect alignments of CSB3-containing reads serves as a proxy for the total number of minicircles that were initially present within the DNA sample. All metrics were processed and visualized using the R function *msc*.*quality* as implemented in the R package rKOMICS (41). In addition, the quality of the assembly was further verified by calling SNPs with BCFtools mpileup/call, retaining only SNPs with QUAL >= 60 and DP >= 30, and assuming that high-quality assemblies should yield relatively low number of homozygous SNPs.

Finally, using the rKOMICS function *msc*.*depth*, minicircle copy numbers (MCN) were estimated as the median read depth per minicircle contig divided by the median genome-wide read depth times two (assuming diploidy in all *Tb* strains).

### Identification of minicircle sequences unique to *Tbg*1

The diversity and similarity of minicircle sequences within all *Tb* strains were examined with the R package rKOMICS (41). Following visual inspection of length distributions using *msc*.*length*, we used *preprocess* to retain minicircle sequences that had the expected length (800-1200 bp) and that were successfully circularized. Retained sequences of all samples were concatenated into a single FASTA file and clustered into Minicircle Sequence Classes (MSCs) based on a minimum percent identity (MPI) of 70, 80, 90 and 95-100 with VSEARCH. In order to choose an appropriate MPI for downstream analyses, we processed VSEARCH clustering results with *msc*.*uc* and inspected - at each MPI - the number of MSCs, the number of perfect alignments and the number of 2-nt and 3-nt gaps. In addition, VSEARCH clustering results were stored into a matrix using the rKOMICS function *msc*.*matrix*, which records the presence (1) or absence (0) of all MSCs (rows) for each strain (columns). This matrix was subsequently used to document the number of MSCs per strain with *msc*.*richness*, to calculate the proportion of MSCs shared between the different *Tb* subspecies with *msc*.*similarity* and to investigate the ancestry among all *Tb* strains with *msc*.*pca*. Finally, we used the rKOMICS function *msc*.*subset* to find MSCs that were present in the *Tbg*1 strains and absent in the strains belonging to the other *Tb* subspecies.

### Development of minicircle-based quantitative PCR assays

For each of the common *Tbg*1-specific MSC, we extracted the assembled minicircle sequences for all 18 *Tbg*1 strains from the alignment using the rKOMICS function *msc*.*seq*, generated consensus sequences with Jalview v2.11.1.4 (52) and designed primers and probes (Table 1) with the RealTimeDesign qPCR assay design software (LGC, Biosearch Technologies). Probes targeting *Tbg*1-specific MSCs were modified with a FAM dye label at the 5’ end and paired with BHQ-1 *plus* at the 3’ end.

**Table 1.**
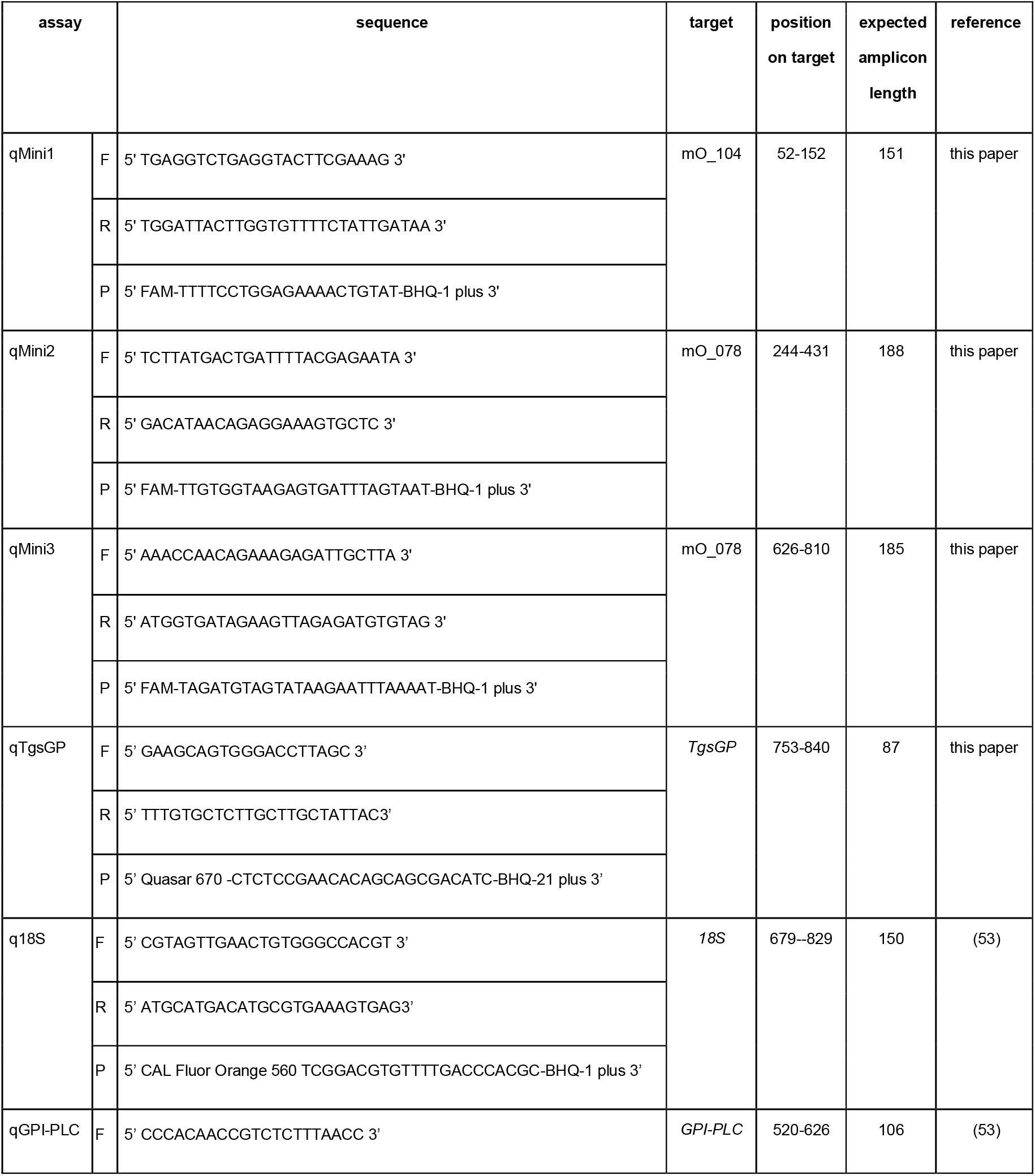
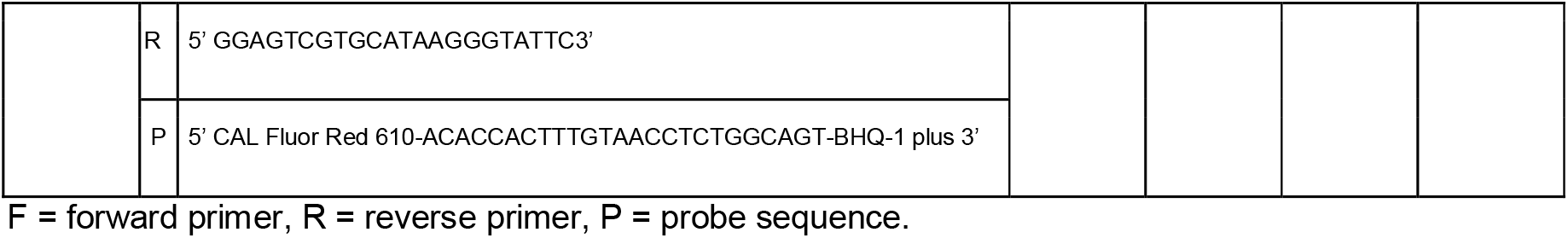
Summary of the various qPCR assays used in this study.

A 20 µl reaction mixture contained 1X PerfeCTa qPCR Toughmix (Quantabio), 100 nM of each primer (LGC, Biosearch Technologies), 300 µM of each probe (LGC, Biosearch Technologies) and 5 µl of template DNA. The thermal cycling profile consisted of an initial denaturation step at 95°C for 10 min followed by 40 cycles at 95°C for 15 s and 60°C for 1 min. qPCR was conducted on a Q-qPCR Instrument (Quantabio), and detection of the quantification cycle (Cq) was calculated using the Q-qPCR Instrument Software v1.0.2 with the automatic threshold enabled.

Each qPCR targeting *Tbg*1-specific MSCs was multiplexed with the *Trypanozoon*-specific q18S-assay targeting the multi-copy 18S rRNA gene (53), the *Trypanozoon*-specific qGPI-PLC-assay targeting the single-copy glycosylphosphatidylinositol-specific phospholipase C (GPI-PLC) gene (53) and a *Tbg*1-specific qTgsGP-assay, designed to target the single-copy TgsGP gene and avoid amplification of TgsGP-like genes (21). The multi-copy 18S rRNA gene was used as an internal standard for the sensitive detection of *Trypanozoon* DNA. The single-copy GPI-PLC gene was used as an internal standard to determine if sufficient *Trypanozoon* DNA was present to detect a single-copy sequence (54), and for the calculation of relative copy numbers (RCN) of each *Tbg*1-specific MSC (see below). The single-copy TgsGP gene was used as a golden standard for the specific detection of *Tbg*1 DNA. The q18S-assay contains a CAL Fluor Orange 56 dye labeled probe paired with BHQ-1 *plus* (53). The qGPI-PLC-assay contains a CAL Fluor Red 610 dye labeled probe paired with BHQ-2 *plus (53)*. The qTgsGP-assay contains a Quasar 670 dye labeled probe paired with BHQ-2 *plus*.

The qPCR efficiency and analytical sensitivity were calculated for each qPCR targeting *Tbg*1-specific MSCs in simplex and in quadruplex format. This was done using phenol:chloroform extracted DNA (see above) of two *Tbg*1 strains. From these DNA extracts, ten-fold serial dilutions in DEPC-Treated Water, ranging from 100 pg/µl to 1 fg/µl, were prepared. Each qPCR was run in quadruplicate for each DNA dilution. A reaction was considered positive if at least three out of four replicates were positive.

The specificity of the quadruplex qPCR assays was assessed with the phenol:chloroform extracted genomic DNA of 34 *Tbb*, 49 *Tbg*1, 7 *Tbg*2, 15 *Tbr*, 2 *T. equiperdum* and 5 *T. evansi* strains (Supplementary Table 2). The 49 *Tbg*1 samples originated from Burkina Faso (2), Cameroon (2), Côte d’Ivoire (6), Congo Brazzaville (3), Democratic Republic of the Congo (35) and South Sudan (1). Note that 26 strains from the Democratic Republic of the Congo were sampled within the context of a treatment outcome study in Mbuji-Mayi (55), with 14 strains sampled from seven patients before (sample name include ‘BT’) and after (‘AT’) treatment. Quadruplex qPCR assays were run in duplicate for each DNA extract. The specificity of the assays was further assessed on DNA prepared from man, cattle, dog, goat, horse, sheep and tsetse (*Glossina fuscipes* from Kwamouth, Democratic Republic of the Congo, 2018), all known hosts of *Tbg*1 (12).

Relative copy numbers (RCN) of each target were calculated using the ΔCq-method with qGPI-PLC as reference. This was done by subtracting the Cq-values obtained for each *Tbg*1-specific MSC from the Cq-value obtained for qGPI-PLC. The resulting ΔCq-value were averaged between replicates and transformed (2^ΔCq) to yield RCNs for each target.

### Annotation of minicircle sequences targeted by qPCR assays

Strain *Tbg*1 340AT (MHOM/CD/INRB/2006/21B) isolated in 2006 in Mbuji-Mayi (DRC) (55) (Supplementary Table 1) was selected for representative minicircle annotation because of its comparatively high minicircle complexity (see results). DNA extraction, sequencing and alignment of sequence reads against the *Tbb* TREU927 v4.6 reference genome was done as described above for the other 38 *Tb* isolates initially included in our study. Reads that did not align to the nuclear reference genome were used for the assembly of mitochondrial maxicircles and minicircles with KOMICS, as described above.

Due to the lack of transcriptomic data for *Tbg*1 strain 340AT, edited mRNA sequences were predicted following a similar approach as in (40). Edited mRNA sequences for *Tbb* strains Lister 427, EATRO 164 and EATRO 1125 (36) were obtained from GenBank and manually corrected for changes in non-T residues based on alignment of the *Tbg*1 340AT maxicircle with the annotated *Tbb* EATRO 1125 maxicircle (36).

Guide RNA prediction and minicircle annotation were performed with python3.7 package *kDNA annotation (36)*. The alignments of the gRNAs encoded on the minicircles targeted by the diagnostic assay to their cognate mRNAs were carefully inspected to identify any ‘non-redundant’ gRNAs, i.e. gRNAs that direct editing events not covered by any other gRNAs. Furthermore, gRNAs 3’ of such non-redundant gRNAs were checked for potential premature truncations by the gRNA calling algorithm by carefully examining the editing capacity of their 3’ end extensions. Any gRNA genes that were confirmed to be non-redundant were considered essential, as were the minicircles that encoded them.

## RESULTS

### Genome analyses confirms the taxon identity of *Tbg*1 strains

The genomes of 38 *Tb* strains were sequenced at a median 159x depth (mean = 155, min = 126, max = 178) (Supplementary Table 3). On average 86.3% of the reads (min = 81.25%, max = 92.05%) aligned to the *Tbb* TREU927 nuclear reference genome (Supplementary Table 3). Initial variant discovery with GATK identified a total of 1,558,963 SNPs across the 38 *Tb* strains. Strict quality filtering and the exclusion of multiallelic sites reduced the data set to 316,287 genome-wide bi-allelic SNPs, of which 310,701 SNPs (98.23%) were located within the 11 megabase chromosomes. In addition, joint genotyping identified a total of 150 SNPs within the maxicircle coding region.

Phylogenetic analyses based on genome-wide SNPs and SNPs from the maxicircle coding region confirmed that all 18 *Tbg*1 parasites clustered together in a monophyletic group, a prerequisite for downstream analyses that aim at identifying *Tbg*1-specific minicircles (Supplementary Figure 1). Using a local BLAST search of assembled contigs, we also confirmed the presence of the *Tbg*1-specific TgsGP gene for the 18 *Tbg*1 strains, and the *Tbr*-specific SRA gene for the six *Tbr* strains. These two genes were absent for the remaining nine *Tbb* strains and five *Tbg*2 strains (Supplementary Table 4).

### Assembly and circularization of kinetoplast minicircles

Mitochondrial minicircles were *de novo* assembled, circularized and reoriented for each of the 38 *Tb* strains using the Python package KOMICS. A total of 9,076 minicircle contigs were assembled across all 38 *Tb* strains, of which 7,156 (78,85%) were successfully circularized (Supplementary Table 5). The length of the majority of circularized minicircles (7,111 contigs, 99.37%) showed a unimodal distribution around ∼1,000 bp (Supplementary Figure 2), which is comparable to the minicircle length found in *Tbb (36)*.

To validate the quality of the assembly process, sequence reads were aligned to the assembled minicircle contigs and several mapping and genotyping statistics were calculated. First, high-quality assemblies should result in relatively low numbers of homozygous SNPs when reads are aligned against the assembled contigs. Here, we identified a total of 302 homozygous SNPs within 127 minicircle contigs (Supplementary Table 5), which is only 1.4% of all assembled minicircle contigs. Only 48 homozygous SNPs were identified within 17 circularized contigs (0.2% of all circularized minicircles) (Supplementary Table 5). These results show that homozygous SNPs were found for only a fraction of the assembled contigs. Second, on average 96.99% of the sequence reads mapped in proper pairs and 94.46% aligned with a mapping quality larger than 20 (Supplementary Table 5), indicating that the large majority of mapped reads aligned with a high quality and with the expected orientation to the minicircle assemblies. Third, we calculated the number of aligned reads containing the CSB3 12-mer as a proxy for the total number of minicircles initially present within the DNA sample. This revealed that on average 93.24% of the CSB3-containing reads aligned against the assembled minicircles, and 88.77% aligned perfectly (Supplementary Table 5), suggesting that we were able to retrieve the vast majority of the minicircles.

### Estimation of maxicircle and minicircle copy numbers and network size

To calculate the average number of maxicircles and minicircles per kinetoplast network, we used the coverage (i.e. median read depth) of the diploid nuclear genome. The average genome-wide coverage was 155 per diploid cell (two copies), and the average coverage per\ haploid sequence was equal to 78 (Supplementary Table 3). The coverage of the kinetoplast maxicircles (coding region only) and minicircles was estimated based on median read depths per 0.1kb. Average coverage of the maxicircle was 1,262 (median = 1,325, min = 99, max = 2,320) (Supplementary Table 6). This equaled an average copy number of 17 maxicircles (median = 18, min = 1, max = 28) per network (Supplementary Table 6), which is slightly lower compared with previous estimates of 20-50 copies per network (36, 56, 57). The average copy number of minicircles (MCN) ranged from 2.1 to 37.9 copies per network in *Tbg*1 strains, from 0.8 to 32.7 copies per network in *Tbb* strains, from 4 to 11.9 copies per network in *Tbg*2 strains and from 3.1 to 11.1 copies per network in *Tbr* strains (Supplementary Table 7). Thus, copy numbers for minicircles within each network varied substantially. The size of each kDNA network was estimated by adding up the estimated copy numbers for all minicircles. These calculations resulted in an estimated average of ∼2,100 minicircles per network, ranging between 252 minicircles in the Nabe strain and 4,828 minicircles in the MSUS-CI-78-TSW-157 strain (Supplementary Table 7). These numbers are slightly lower compared to earlier estimates of 5,000-10,000 minicircles per network (36, 56–58).

### Sequence diversity and similarity of kinetoplast minicircles

Minicircle sequence diversity was examined using a clustering approach, whereby minicircles were grouped into MSCs based on a minimum percent identity. This was done on the 7,111 circularized minicircle contigs of the expected length, as these would produce the most robust alignments. At 100% identity, a total of 5,883 unique MSCs were identified across the 38 *Tb* strains, leaving 1,228 MSCs that are shared between two or more isolates.

The number of MSCs decreased sharply with decreasing percent identities to a total of 719 MSCs at 70% identity (Supplementary Figure 3A). Regardless of the percent identity used, *Tbg*1 parasites contained an average of 103 MSCs per strain (median = 107), which is on average 2.48-fold lower compared to other taxa of the *Trypanozoon* subgenus (Figure 1). Most of the *Tbg*1 strains displayed a fairly similar number of MSCs, ranging between 89 and 122 MSCs (Figure 1), with the exception of LiTat 1.5 (50 MSCs) and Bosendja (76 MSCs). The composition of minicircle sequences was further investigated by quantifying the proportion of MSCs unique to *Tbg*1. At 98%-100% identity, the 18 *Tbg*1 parasites did not share any MSC with the non-*Tbg*1 subspecies (*Tbr, Tbg*2 and *Tbb*) (Supplementary Figure 3B). Below 98%, there was a steady increase in the proportion of shared MSCs between *Tbg*1 and non-*Tbg*1 subspecies (Supplementary Figure 3B). At the 98% identity threshold, the *Tbg*1 group contained 241 MSCs, none of which were found in the other *Tb* subspecies and three of which were found in all 18 *Tbg*1 strains (Figure 2). These three *Tbg*1-specific MSCs were retained as candidate markers for our new molecular test.

**Figure 1.**
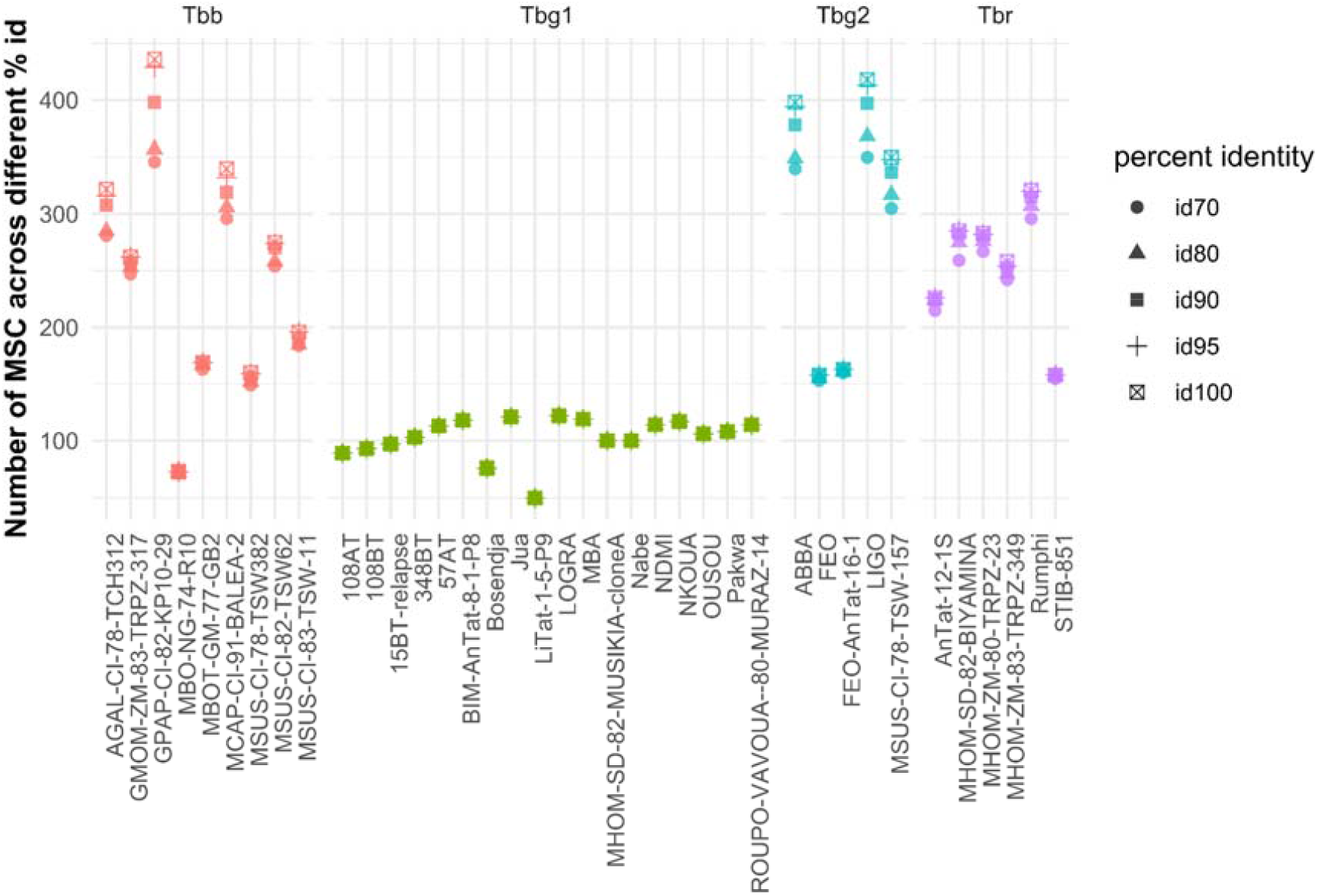
Number of Minicircle Sequence Classes (MSCs) for 38 *Tb* strains. Following *de novo* assembly and circularization, mitochondrial minicircle sequences were clustered into groups of sequences sharing a minimum percent identity (MSCs). Here, we summarized the number of MSCs identified within each *Tb* strain for a range of percent identities (70, 80, 90, 95 and 100). Strains were grouped as *T. b. brucei* (*Tbb*), *T. b. gambiense* type 1 (*Tbg*1), *T. b. gambiense* type 2 (*Tbg*2) and *T. b. rhodesiense* (*Tbr*).

**Figure 2.**
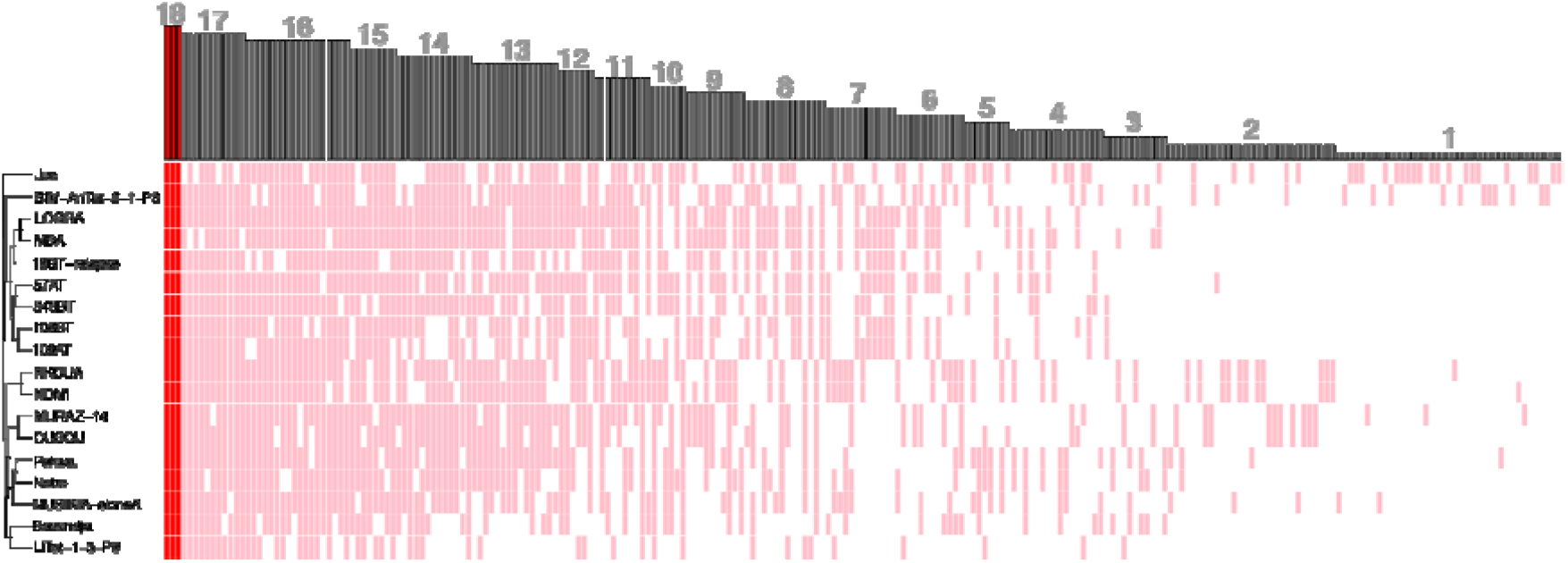
Minicircle sequence diversity in 18 *Tbg*1 strains. Clustering at the 98% identity threshold revealed a total of 241 MSCs that were present in at least one of 18 *Tbg*1 strains and absent in the *Tbb, Tbr* and *Tbg2* strains. Heatmap summarizes the presence/absence of these 241 MSCs (columns) in the 18 *Tbg1* strains (rows). Gray lines indicate the presence of a given MSC in a given *Tbg*1 strain. Dark red lines indicate the three MSCs that were found in all 18 *Tbg*1 strains. Barplot on top of the plot shows the number of strains harboring a given MSC.

### Novel multiplex qPCRs including *Tbg*1-specific kinetoplast minicircles as target

Using the RealTimeDesign qPCR assay design software, we successfully designed three simplex qPCR assays (here-after referred to as qMini1, qMini2 and qMini3) targeting two *Tbg*1*-*specific MSCs (here-after referred to as mO_078 and mO_104) (Table 1). Due to limitations such as ambiguous base count, low GC percentage and low melting temperatures, we were unable to design primers and probes for the third *Tbg*1*-*specific MSC. The *Tbg*1 specificity of the designed primers and probes was confirmed with a BLAST search (48) on TriTrypDB (https://tritrypdb.org), with a Primer BLAST search on NCBI (https://www.ncbi.nlm.nih.gov) and with the command line search tool grep in sequencing reads generated by this study. The three simplex qPCR assays qMini1, qMini2 and qMini3 were each multiplexed with q18S, qGPI-PLC and qTgsGP (Table 1) to produce three quadruplex reactions (here-after referred to as *g*-qPCR1, *g*-qPCR2 and *g*-qPCR3 when qMini1, qMini2 and qMini3 were included, respectively).

The efficiency and analytical sensitivity of the simplex and quadruplex qPCR assays were investigated using DNA from two *Tbg1* strains (Supplementary Table 8), one with relatively low MCNs (Nabe; average MCN = 1) and one with relatively high MCNs (LOGRA; average MCN = 8). The lower detection limit of 0.05 pg DNA was reached in both strains for qMini1, qMini2 and qMini3 in their respective simplex reactions (Supplementary Figure 4), for q18S in all three *g*-qPCR assays (Supplementary Figure 4), and for qMini1 and qMini3 in their respective *g*-qPCR assays, with the exception of qMini3 that achieved the detection limit of 0.5 pg DNA in the Nabe strain (Supplementary Figure 4). This detection limit of 0.5 pg DNA was also reached by qTgsGP and/or qGPI-PLC in all three *g*-qPCR assays with the LOGRA strain and in the *g*-qPCR2 and *g*-qPCR3 assays with the Nabe strain (Supplementary Figure 4). The analytical sensitivity of qMini2 was greatly reduced in the *g*-qPCR2 assay to 100 pg DNA with the Nabe strain and 0.5 pg DNA with the LOGRA strain (Supplementary Figure 4). The qPCR efficiency of *g*-qPCR1 and g-qPCR3 was estimated between 93% and 106% (Supplementary Figure 4), which is considered acceptable (https://www.thermofisher.com/content/dam/LifeTech/global/Forms/PDF/real-time-pcr-handbook.pdf).

### The qMini3 assay displays a similar specificity as the qTgsGP assay

To assess the taxon-specificity of qMini1 and qMini3 within the *Trypanozoon* subgenus, a total of 118 DNA extracts were tested with the *g*-qPCR1 and *g*-qPCR3 assays (Supplementary Table 9). Here, qMini2 was excluded because of its low analytical sensitivity in the *g*-qPCR2 assay (see above) and because it targets the same minicircle as qMini3 (Table 1). Six of the 118 DNA extracts were excluded as they didn’t react in duplicate with qGPI-PLC, which was used as an internal standard to determine if sufficient *Trypanozoon* DNA was present to detect a single-copy sequence (Supplementary Table 10). The remaining 112 DNA extracts reacted with the *Trypanozoon*-specific assays q18S and qGPI-PLC (Supplementary Table 9). All 49 *Tbg*1 DNA extracts reacted with qTgsGP, qMini1 and qMini3, with the exception of MSUS/CI/82/TSW125_KP1_cloneB (*Tbg*1 isolated from a pig in Côte d’Ivoire), ALJO (*Tbg*1 isolated from a human patient in DRC) and GUIWI-BOBO80-MURAZ18 (*Tbg*1 isolated from a patient in Burkina Faso) that remained negative for qMini1. The qMini1 assay also showed one cross reaction with DNA extracted from the *Tbr* strain Etat 1.2 R, with a Cq-value 5x lower than that of the qGPI-PLC. The qMini3 assay showed no cross reactions or false negative results. In addition, the *g*-qPCR3 assay did not amplify DNA from human and non-human vertebrates that are known to be susceptible for *Tbg*1 infection, *in casu* horse, cattle, goat, sheep, pig and dog, and DNA from *Glossina fuscipe*s. Also, DNA from other livestock affecting trypanosomes like *T. congolense, T. theileri* and *T. vivax* was not amplified (Supplementary Table 9).

### The qMini3 assay targets a minicircle with a relatively high, but variable copy number

Relative Copy Numbers (RCN) of the minicircle sequence targeted by qMini3 (RCN_mO_078_), the target sequence of qTgsGP (RCN_qTgsGP_) and the target sequence of q18S (RCN_q18S_) were calculated for each of the 49 *Tbg*1 DNA extracts using the ΔCq-method (Supplementary Table 11). The median RCN_mO_078_ (7.98) was 4.29x higher than the median RCN_qTgsGP_ (1.86) and 2.06x higher than the median RCN_q18S_ (3.88) (Figure 3A). However, RCN_mO_078_ also displayed a larger variation (SD = 9.01, min = 0.77, max = 40.58) compared to RCN_qTgsGP_ (SD = 0.62, min = 0.94, max = 3.47) and RCN_q18S_ (SD = 0.70, min = 1.51, max = 5.47). The RCN_mO_078_ was lower compared to RCN_qTgsGP_ for 4/49 *Tbg*1 strains and to RCN_q18S_ for 12/49 *Tbg*1 strains. There was no association between RCN_mO_078_ with the year of isolation (Pearson correlation test; cor= -0.1925748, t = -1.331, df = 46, p-value = 0.1897) or the geographical origin of the strain (Kruskal-Wallis test; country: chi-squared = 4.0896, df = 5, *p*-value = 0.5366; region: chi-squared = 3.0453, df = 2, *p*-value = 0.2181). High variation in RCN_mO_078_ was found within one group of *Tbg*1 strains isolated from humans between 2005 and 2009 in Mbuji-Mayi (Democratic Republic of the Congo), with a minimum RCN_mO_078_ of 0.8 in the 186BT strain and a maximum RCN_mO_078_ of 36.7 in the 93AT strain. Here, RCN_mO_078_ was significantly higher in strains sampled after treatment (mean RCN_mO_078_ = 14.9) compared to the RCN_mO_078_ in strains sampled before treatment (mean RCN_mO_078_ = 6.9), although this difference was not significant (Welch two sample t-test on all strains, t=2.0305, df = 11.944, *p*=0.06519 and paired t-test on paired strains, t=1.5476, df=5, *p*=0.183). The RCN_mO_078_ as calculated using the ΔCq-method was strongly associated with MCN_mO_078_ as calculated using standardized read depths, with a coefficient of determination of 0.91, a slope of 0.32 and a y-intercept of 0.10 (Figure 3B).

**Figure 3.**
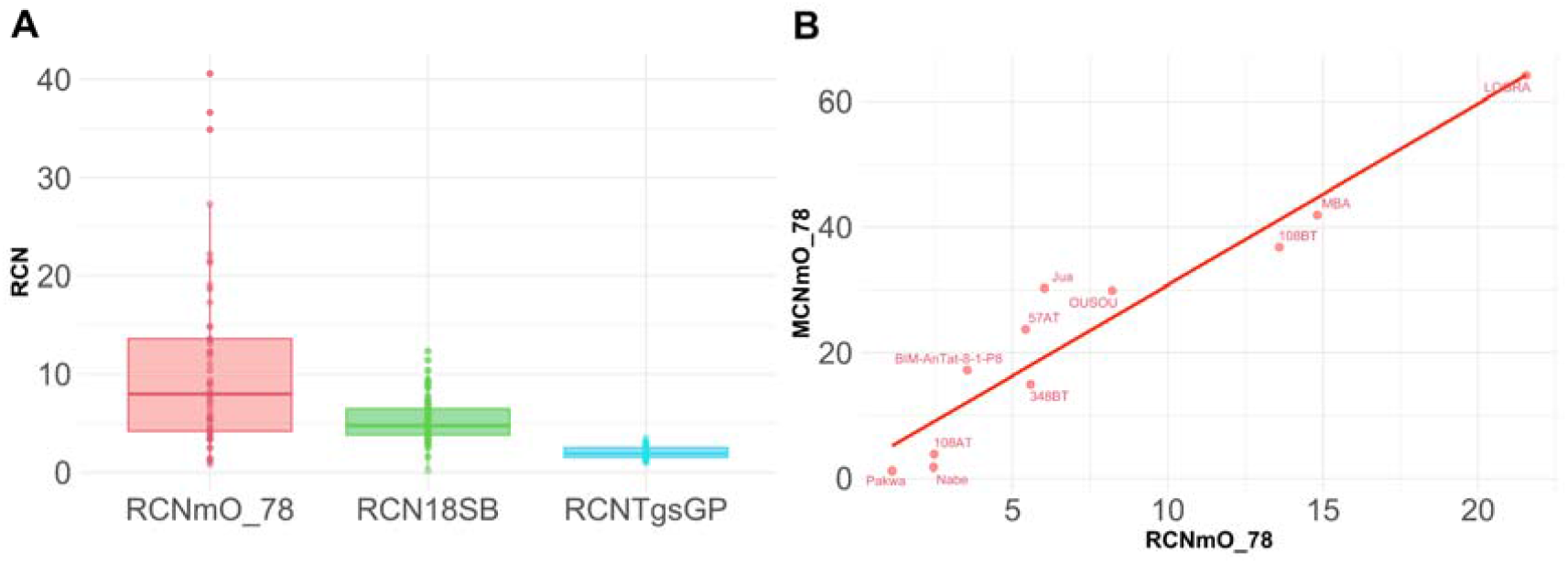
Relative Copy Numbers of the qMini3 target sequence. **(A)** Relative copy numbers (RCN) were estimated for the qMini3, q18S and qTgsGP target sequences using the ΔCq-method with the qGPI-PLC as reference. Boxplots summarize the RCN estimates as calculated for 49 *Tbg*1 strains. **(B)** Scatter plot showing the relationship between RCN (x-axis) and MCN (y-axis). To test whether RCN as calculated using the ΔCq-method is comparable to the Minicircle Copy Numbers (MCN) as calculated using standardized read depths, RCN and MCN were calculated for the qMini3 target sequence for 11 out of 18 *Tbg*1 strains. For these 11 strains, there was sufficient DNA to allow both whole genome sequencing (for MCN calculation) and a qPCR run (for RCN calculation) on the same DNA extract. The remaining seven strains were excluded here as there wasn’t sufficient DNA left for a qPCR run following whole genome sequencing.

### The qMini3 assay targets a minicircle containing non-redundant guide RNA genes

Annotation of minicircles was done for strain 340AT. A local BLAST search of assembled contigs revealed the presence of the *Tbg*1-specific *TgsGP* gene, confirming that 340AT is a *Tbg*1 strain (Supplementary Table 3). The kinetoplast maxicircle and minicircles were assembled with KOMICS using sequence reads that did not align to the nuclear reference genome. This resulted in a maxicicle contig of 21,287bp long (including the entire coding region) and a total of 143 minicircle contigs (including 129 circularized contigs). Hence, 340AT has the highest number of minicircles when compared to the 18 *Tbg*1 strains (max. 132 minicircles) initially sequenced in this study (Supplementary Table 7), which was the main motivation for using the 340AT data for representative minicircle annotation.

Annotation of the 143 minicircles revealed that the minicircle targeted by qMini3 encodes four gRNA genes (Figure 4). These gRNAs are involved in editing of the maxicircle genes cytochrome c oxidase subunit 3 (gCOX3(616-656) and gCOX3(341-369)), ATPase subunit 6 (gA6(415-452)) and NADH dehydrogenase subunit 7 (gND7(847-888)) (Figure 4). As expected, the minicircles contain the semi-conserved region characterized by conserved sequence blocks CSB1, CSB2 and CSB3 (36, 50), and the gRNA genes are framed by imperfect 18bp inverted repeats (36, 59–61).

**Figure 4.**
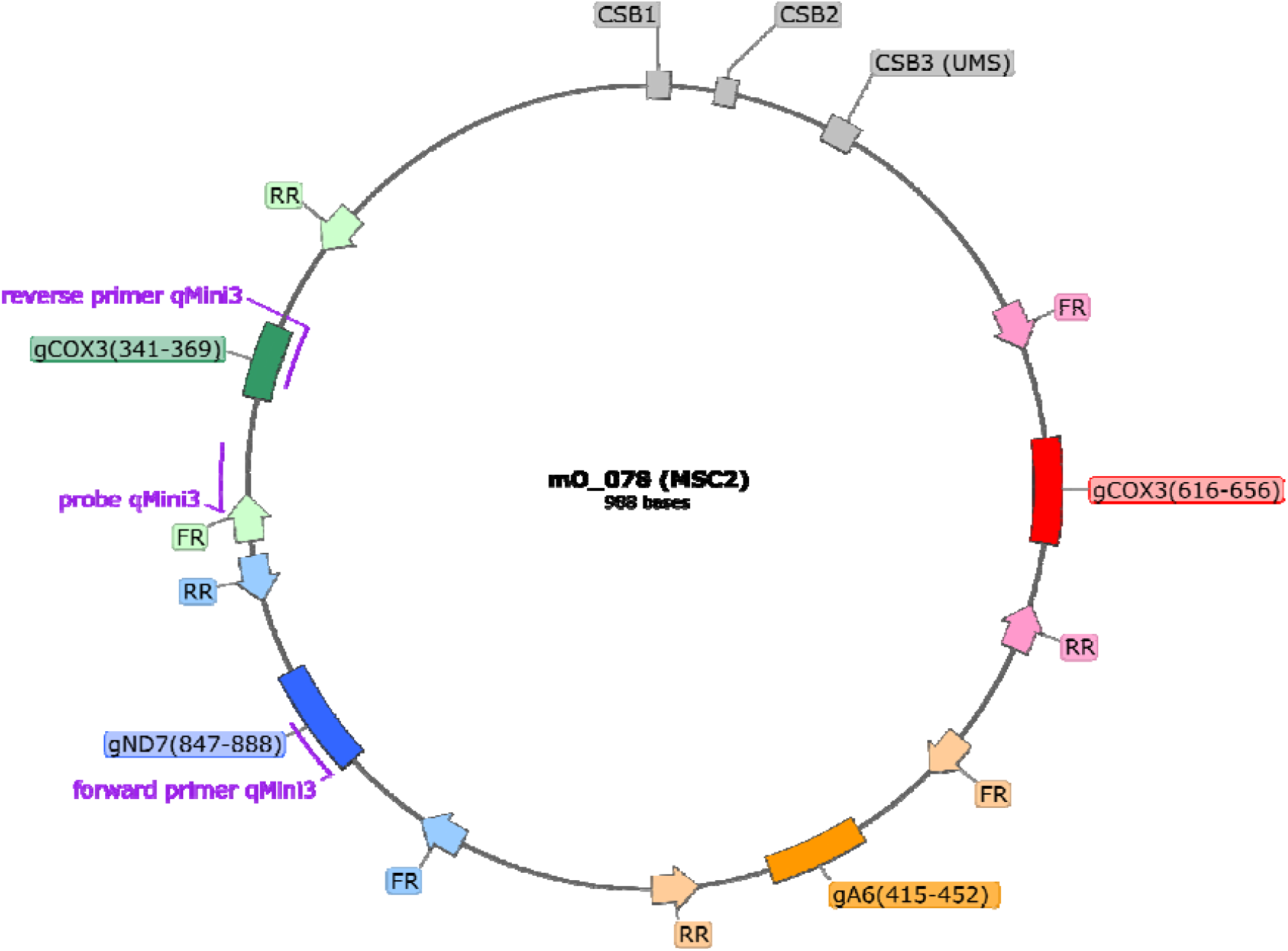
Annotation of minicircle mO_78 targeted by qMini3. Binding sites for the diagnostic PCR primer pair and the corresponding probe for qMini3 are indicated with purple lines. Conserved sequence blocks CSB1, CSB2 and CSB3 (or universal minicircle sequence, UMS) (gray boxes), the four encoded gRNA genes (red, orange, blue and green boxes), and the 18-bp inverted repeats (block arrows) that flank the gRNA genes are also indicated. This figure was generated with SnapGene (http://www.snapgene.com).

Next, we investigated if any of the gRNAs encoded by minicircle mO_078 are non-redundant, i.e. whether they direct the editing of sites not covered by any of the other gRNAs encoded in this strain’s kDNA. Minicircles encoding only redundant gRNAs might be more prone to loss due to lack of selective pressure, which could make a diagnostic assay based on such minicircles less reliable. Our analyses showed that gRNAs gA6(415-452) and gND7(847-888) direct editing events not covered by any other gRNA (Figure 5), and are thus non-redundant. The two COX3 gRNAs are redundant with gRNAs encoded by other minicircles (results not shown).

**Figure 5.**
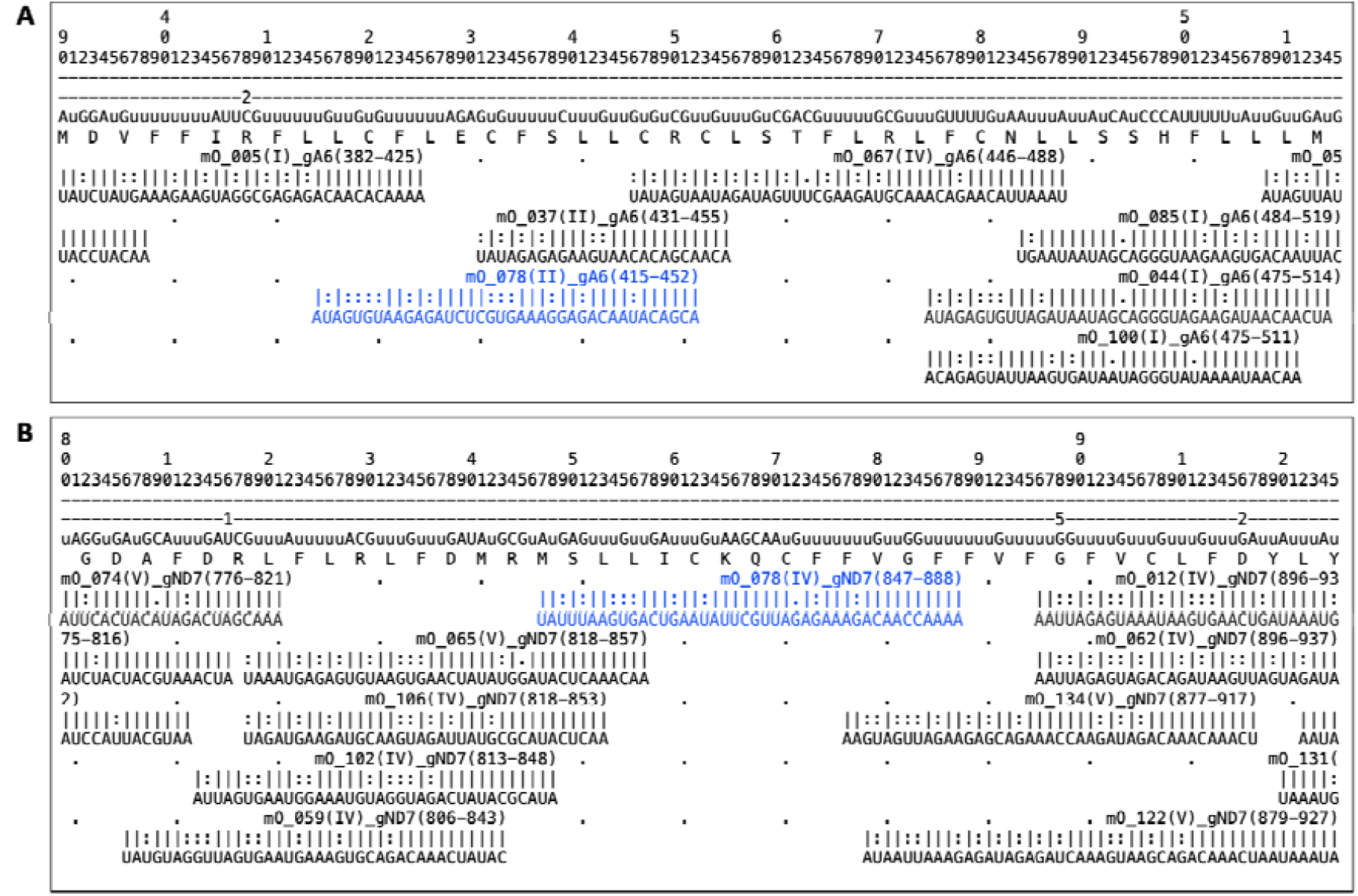
Alignment of the non-redundant gRNAs from the qMini3 targeted minicircle mO_078 to their target mRNAs. The alignment shows the non-redundant gRNAs in blue (**A**, gA6(415-452); **B**, gND7(847-888)) as well as the mRNA sequences immediately upstream and downstream, along with the neighbouring gRNAs encoded by other minicircles. Lines 1-3: mRNA position (hundreds, tens, ones); line 4: number of Us that have been deleted from the pre-edited mRNA at this position; line 5: edited mRNA sequence 5’ to 3’ (lowercase “u”s represent insertions). Editing events directed by the non-redundant gRNA are shown in red. Note that the ‘anchor’ sequence at the 5’ end of each gRNA cannot direct editing events; line 6: protein sequence. For each gRNA: Line 1: name (mO_name(cassette position)_mRNA(start-end of alignment on mRNA)), underscore characters denote the anchor; line 2: base-pairing: “|”: Watson-Crick basepair, “:”: GU basepair, “.”: mismatch basepair; line 3: gRNA sequence 3’ to 5’.

## DISCUSSION

This study presents a computational investigation of mitochondrial minicircle sequence diversity in trypanosome isolates, resulting in the development of a qPCR assay as a promising new tool for sensitive diagnosis of *Tbg*1 infections in humans and animals.

Computational and phylogenetic analyses using genome sequencing data revealed that the 18 sequenced *Tbg*1 strains are monophyletic and contain the *TgsGP* gene, an essential precondition for identifying *Tbg*1-specific minicircles. The latter was achieved by grouping *de novo* assembled and circularized minicircles into MSCs according to sequence similarity. This uncovered a variable number of MSCs within the *Trypanozoon* subgenus, with a considerably lower number of MSCs in *Tbg*1 compared to the other subspecies. The comparatively lower number of MSC within *Tbg*1 may be the result of its asexual evolution (5) that results in the inevitable loss of redundant minicircles due to random genetic drift (62).

Hence, the monophyletic origin and the asexual evolution of *Tbg*1 may explain the less complex minicircle populations in isolates for this subspecies, with conserved presence of some minicircle classes, which facilitated the identification of *Tbg*1-specific minicircles. Identification of taxon-specific minicircles may prove more challenging for trypanosomatid parasites experiencing occasional recombination (63–65), leading to heterogeneous minicircle populations as a result of biparental inheritance of mitochondrial minicircles (40, 66–68).

A total of 241 MSCs were recognized as *Tbg*1-specific, of which three were shared by the 18 studied *Tbg*1 strains. For two of the three *Tbg*1-specific minicircles (mO_078 and mO_104), three molecular assays could be successfully developed and tested. While two qPCR assays were discontinued because of false-negative results or low analytical sensitivities, one qPCR assay targeting minicircle mO_078 was fully specific (i.e. no false positives or false negatives) when tested on DNA of 112 different *Trypanosoma sp*. strains. These results show that the minicircle-based assay is as specific as the assay targeting the *TgsGP* gene, the current golden standard for molecular detection of *Tbg*1 parasites (69–75), confirming the taxon-specificity of some kDNA minicircles in *T*.*b. gambiense* (33, 34) and their exploitability in molecular tests as has been described for *Leishmania* (37, 38) and *T. evansi* type A and B (39, 76, 77). Annotation of the mO_078 minicircle demonstrated that it encodes two non-redundant gRNAs that are essential for completing the editing of ATP synthase subunit A6, a gene required for survival in both the bloodstream stage and the insect stage of *Tb* (78, 79), and NADH dehydrogenase subunit 7, respectively. Hence, our results indicate that minicircle mO_078 is essential for the survival of the parasite and will most likely be preserved in all natural *Tbg*1 strains, ensuring the reliability of a diagnostic assay targeting this minicircle.

Molecular analyses revealed that the *Tbg*1-specific minicircle mO_078 is a multicopy marker for the large majority of strains tested in this study, with a median copy number equal to eight and a maximum copy number of 41. Minicircle mO_078 copy number exceeded that of the multicopy *18S* gene in ¾ of the *Tbg*1 strains and was as low as the *TgsGP* copy number in only four of the 49 tested *Tbg*1 strains. This finding confirmed the multicopy nature of mitochondrial minicircles in *Trypanosoma* and *Leishmania* parasites (36, 80). However, our results also indicated that there is a relatively large variation in minicircle copy numbers across strains, suggesting that the detection limit of the minicircle-based assay may depend on the strain being investigated. The detection limit may also depend on the DNA extraction method, as DNA-extraction based on spin columns causes a random loss of small molecules like minicircles (36). Here, we avoided such biases by using the phenol-chloroform extraction method that captures all nucleic acids, although this method may be less amenable for high-throughput processing of human and animal specimens.

The high specificity and generally high copy number of minicircle mO_078 makes this a promising new marker for sensitive diagnosis of *Tbg*1 infections. Specifically, within the context of reaching EOT of *g*HAT by 2030 (9), our minicircle-based assay may prove valuable for studying the role of an animal reservoir in the epidemiology of *g*HAT (12).

Therefore, we proposed a new multiplex qPCR assay (*g*-qPCR3) that targets the *Tbg*1-specific minicircle mO_078 (serving as sensitive detection of *Tbg*1) in combination with the *Trypanozoon*-specific *18S* (serving as sensitive detection of *Tb s*.*l*.) and the *Tbg*1-specific *TgsGP* gene (which confirms the trypanosome is human-infective and thus of epidemiological significance). We have shown that the *g*-qPCR3 assay is applicable on animals, as it does not amplify DNA from other livestock affecting trypanosomes like *T. congolense, T. theileri* and *T. vivax*, from the tsetse fly *Glossina fuscipe*s and from six livestock species that are known to be susceptible to *Tbg*1 infection. Based on these data, we also trust that the specificity will not be compromised when testing blood from wild *Bovidae* and non-human primates (12), although we had no access to specimens from wild fauna to formally test this. The main limitation of our study is that the *g*-qPCR3 assay was tested on only one *Tbg*1 strain isolated from a host other than human, *in casu* a pig from Côte d’Ivoire (81, 82), mainly because of the scarcity of such samples. However, given that minicircle mO_078 is non-redundant and universal (computational analyses confirmed that mO_078 was present in 193 sequenced *Tbg*1 strains sampled from humans in 11 different countries (data not shown)), we are confident that the *g*-qPCR3 assay will be successful at amplifying *Tbg*1 DNA from livestock specimens.

In conclusion, this study exemplifies the power of genome assembly and annotation for identifying species-specific multicopy genetic markers. We developed a minicircle-based assay that is as specific as the current golden standard for molecular detection of *Tbg*1 infections, and argued that the *g*-qPCR3 has the diagnostic potential for assessing the importance of an animal reservoir in the epidemiology of *g*HAT.

## Data Availability

Sequence reads generated within the context of this study have been deposited in the European Nucleotide Archive under accession number PRJEB49966 (https://www.ebi.ac.uk/ena/browser/view/PRJEB49966). The sequence of the Tbg1-specific minicircle mO_078 was deposited to NCBI under accession number OM238297. Scripts used for processing WGS data in this study are available at https://github.com/GeertsManon/g-qPCR. All graphical analyses were performed using R v4.0.3 in RStudio v1.4.1103.

## FUNDING

PB received financial support from the Bill & Melinda Gates Foundation (grant number OPP1174221) and the Flemish Government EWI SOFI-2018 “Cryptic human and animal reservoirs compromise the sustained elimination of gambiense-human African trypanosomiasis in the Democratic Republic of the Congo”. FVdB was supported by the Department of Economy, Science and Innovation in Flanders and by the Research Foundation Flanders (Grants 1226120N and 1528117N). AS is supported by the UK Medical Research Council Fellowship MR/L019701/1. ZC is supported by an EASTBIO PhD studentship from the UK Biotechnology and Biological Sciences Research Council.

## ACKNOWLEDGEMENTS

We thank Prof. Van Den Abbeele for critical reading of the manuscript, Isabel Saldanha and Steve Torr from the Liverpool School of Tropical Medicine for providing tsetse fly DNA and Dr. Vet. Xanthe Helsen and Dr. Vet. Wauter Van Deun for providing us with cattle, dog, goat, horse, pig and sheep blood.

## DATA AND SCRIPT AVAILABILITY

Sequence reads generated within the context of this study have been deposited in the European Nucleotide Archive under accession number PRJEB49966 (https://www.ebi.ac.uk/ena/browser/view/PRJEB49966). The sequence of the *Tbg*1-specific minicircle mO_078 was deposited to NCBI under accession number OM238297. Scripts used for processing WGS data in this study are available at https://github.com/GeertsManon/g-qPCR. All graphical analyses were performed using R v4.0.3 in RStudio v1.4.1103.

## Notes

### Competing Interest Statement

The authors have declared no competing interest.

