## Supplementary Figures for "Deep kinetoplast genome analyses result in a novel molecular assay for detecting *Trypanosoma brucei gambiense*-specific minicircles"

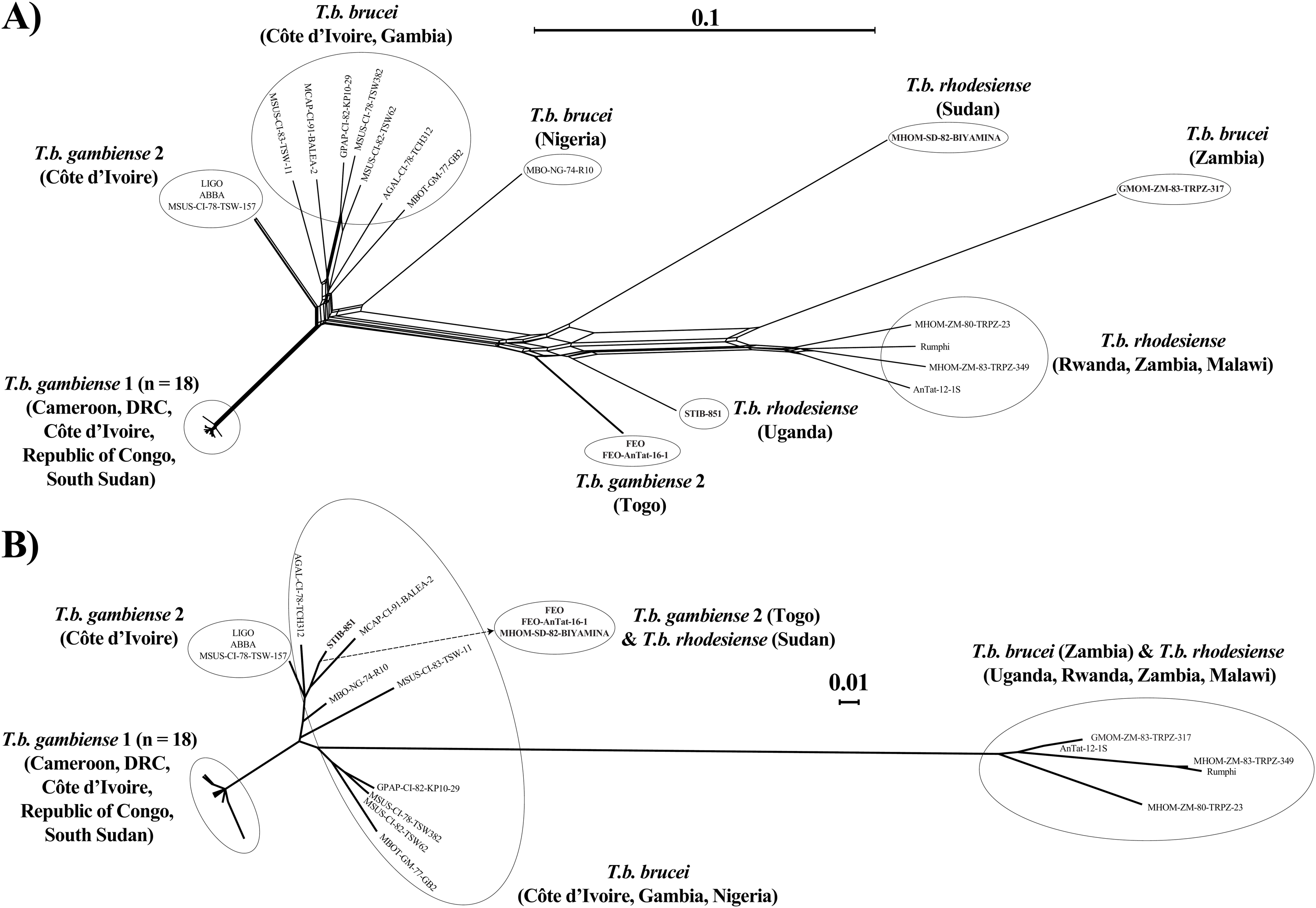

**Supplementary Figure 1.** **(A)** Phylogenetic network based on uncorrected *p*-distances as estimated with 316,287 genome-wide SNPs. **(B)** Neighbour-Joining phylogenetic tree based on uncorrected *p*-distances as estimated with 150 SNPs from the mitochondrial maxicircle coding region.

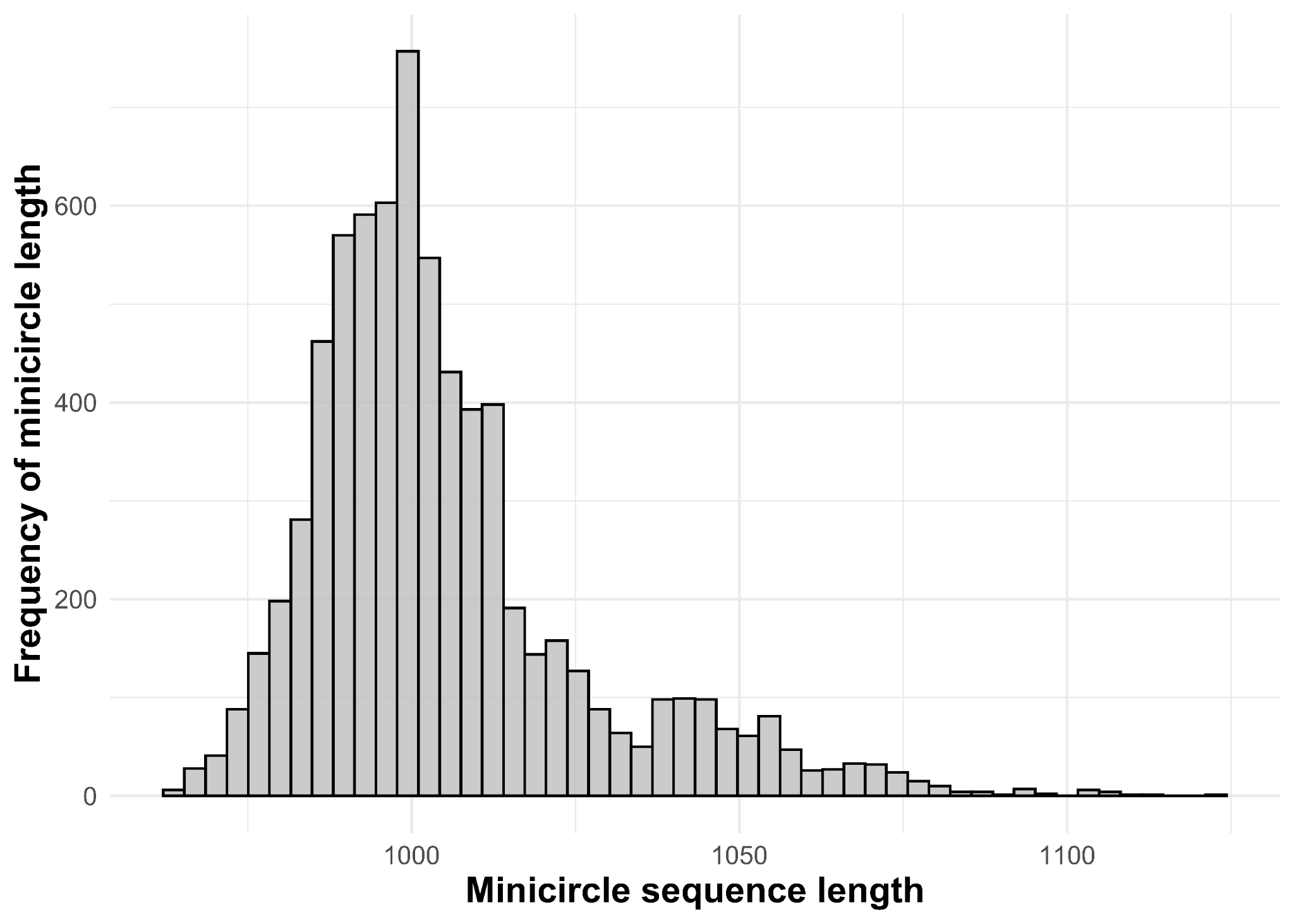

**Supplementary Figure 2.** Length in base pairs (bp) of the 7,111 circularized minicircle contigs of expected length (~1,000 bp) in 38 *Tb* strains.

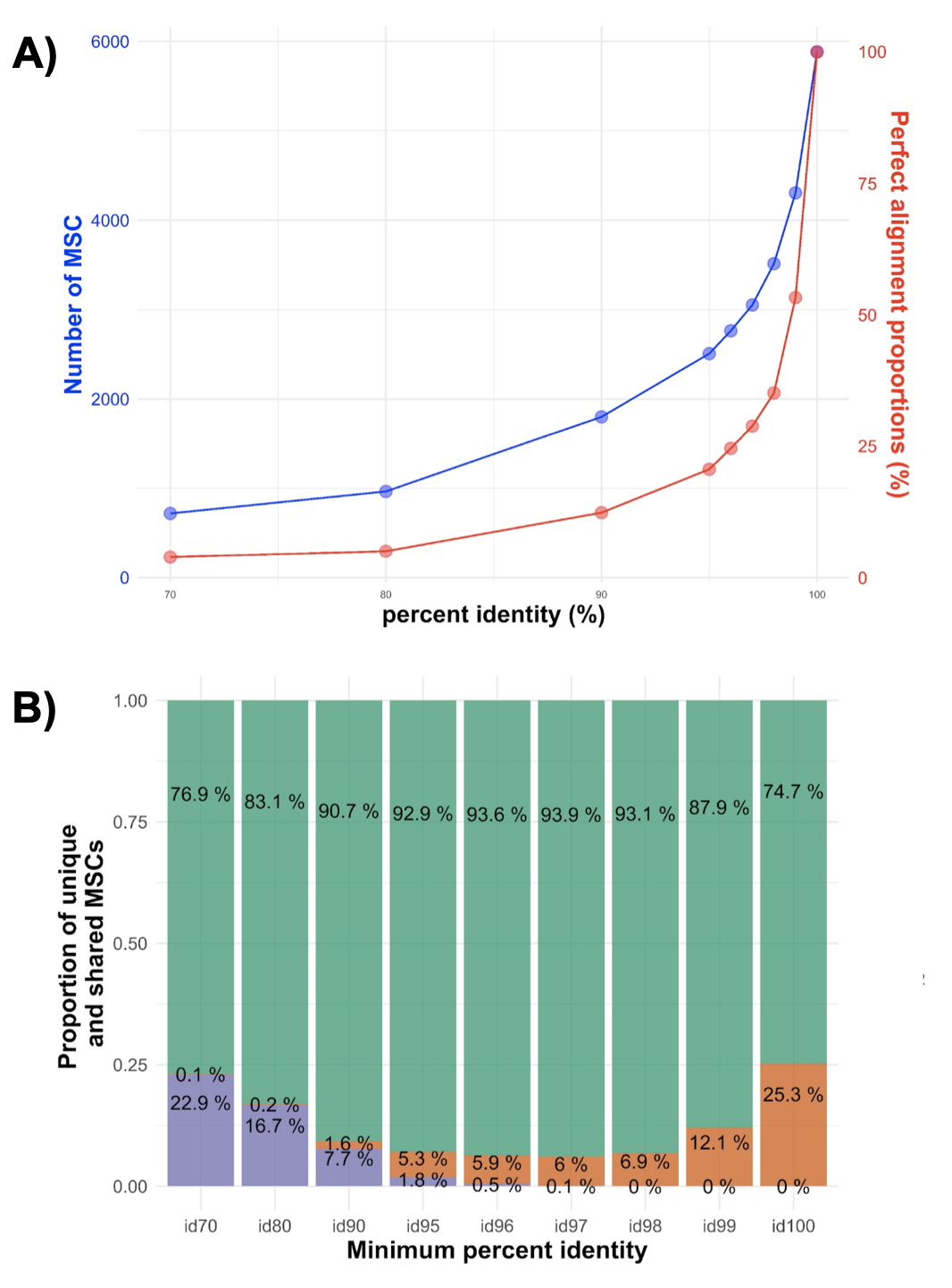

**Supplementary Figure 3.** **(A)** Clustering results of the 7,111 circularized minicircle contigs, showing the number of Minicircle Sequence Classes (MSC) and proportion of perfect alignments (i.e. proportion of alignments without any gaps) for each percent identity. **(B)** Proportion of MSCs that are unique to *Tbg*1 (orange), unique to non-*Tbg*1 parasites (green) and that are shared between between *Tbg*1 and non-*Tbg*1 parasites (purple) for each percent identity.

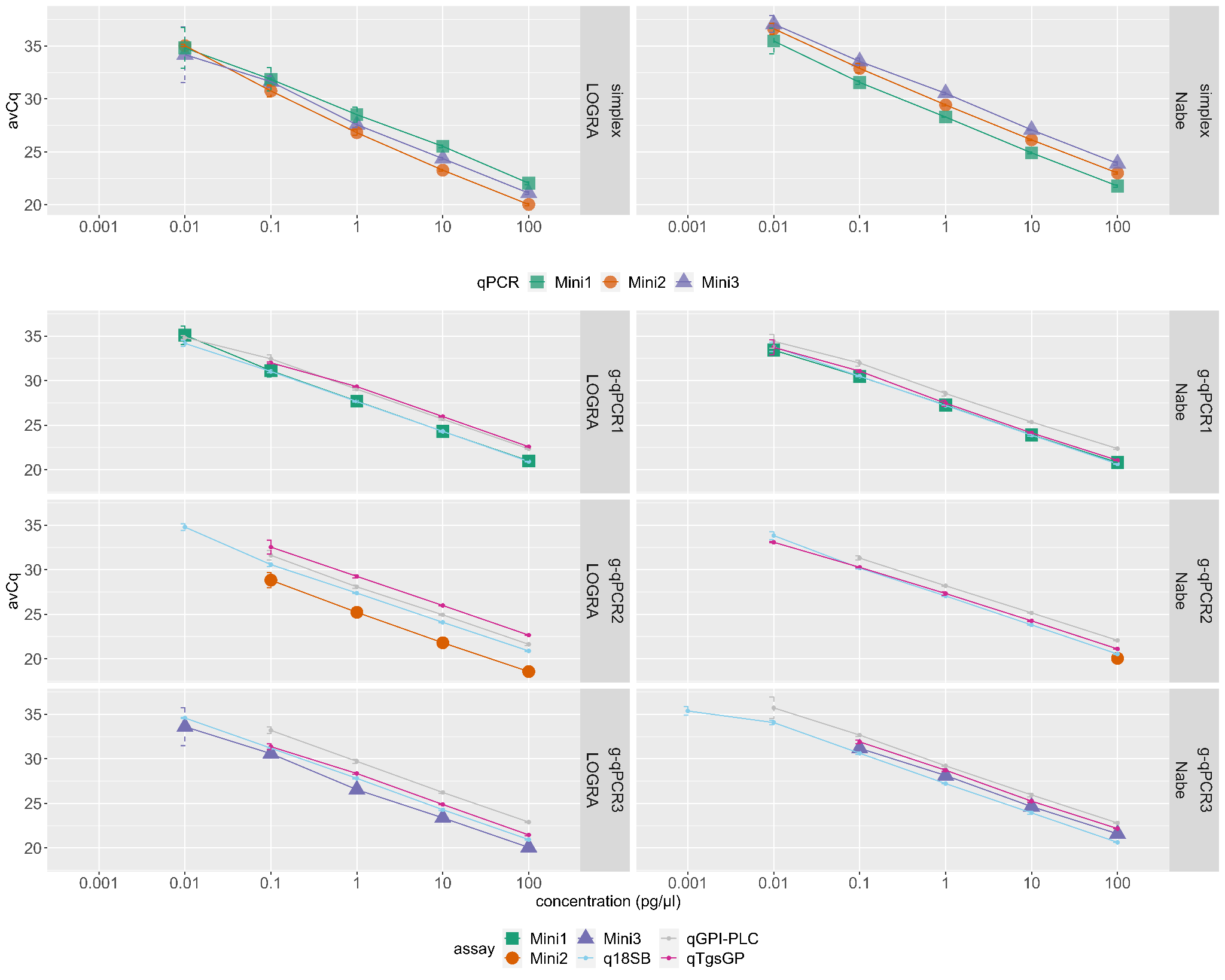
**Supplementary Figure 4.** qPCR efficiencies of the novel qPCRs targeting *Tbg*1-specific MSCs in simplex and quadruplex format. Cq-values were obtained from tenfold dilution series, ranging from 100 pg/µl down to 0.001, on DNA of two Tbg1 strains: one with relatively low (Nabe, bottom) and one with relatively high MCNs (LOGRA, top). Mean and standard deviation were calculated when at least three out of four replicates showed amplification. A linear regression was fitted to calculate the slopes and the qPCR efficiencies.
